# Associations of schooling type, qualification type, and subsequent health in mid-adulthood: Evidence from the 1970 British Cohort Study

**DOI:** 10.1101/2023.07.12.23292261

**Authors:** Keyao Deng, Liam Wright, Richard Silverwood, Alice Sullivan, David Bann

## Abstract

**Background:** Education is thought to benefit subsequent health. However, existing studies have predominantly focused on educational attainment while the type of educational institution attended has been overlooked, despite being important indictors of education resource, quality, and influencing subsequent socioeconomic outcomes. In this study, we investigated associations between type of high school or university attended and multiple subsequent health outcomes in mid-adulthood.

**Methods:** Data from the 1970 British Cohort Study were used (N=8 107). Type of high school attended (comprehensive; grammar; private) was ascertained at age 16 and type of university (classified as higher (Russell Group) or normal-status) was reported at age 42. We investigated ten health measures that capture cardiometabolic risks, physical capabilities, and cognitive function measured at age 46. Multivariable regression models were used, adjusting for sex and childhood socioeconomic, health and cognitive factors.

**Results:** In unadjusted models, private school and higher-status university attendance were favourably associated with most health outcomes. After adjusting for potential confounders, associations between private school attendance and cardiometabolic risks remained; associations for higher-status university attendance and cognitive function remained, while the association with other health outcomes were largely attenuated. For example, private school attendance was associated with 0.14 standard deviation (SD) (95% confidence interval (CI): 0.04, 0.23) lower body mass index (BMI) while higher-status university attendance was associated with a 0.16 SD (0.07, 0.26) better performance in memory (word list recall task) compared with normal-status university attendance.

**Conclusions:** The type of educational institution attended was associated with multiple health outcomes—it could therefore be an important under-researched component of health inequalities. Further research is warranted to test the causal nature of this relationship and its generalisability to more recently born cohorts given changes in secondary education and the expansion of higher education.

## Introduction

A well-documented relationship between education and health exists, with people of higher education typically having lower mortality and morbidity rates compared with their less educated counterparts (1,2). Education is associated with a wide range of physical and mental health outcomes (3–6), with some evidence that such links are causal in nature (7–9). However, most studies investigating this education-health association have focused solely on educational attainment, ignoring other aspects of education.

The type of education (i.e., attending a higher-status institution as opposed to a normal-status institution) an individual attends tends to affect both educational and other later-life outcomes, in particular earnings and occupation (10). In the UK, the schooling system consists of two types of schools: private (independent) and public (state-funded). The private schools are characterised by having minimal state involvement, more resourced and being highly socially and economically selective (11). Only around 7 percent of British students are attending private schools and the participation is concentrated at the top of the family income distribution (12). Private school attendance was found to be associated with a greater economic premium in the labour market, with privately educated workers more likely to enter occupations requiring greater leadership skills (11).

Several potential mechanisms may link the type of education and subsequent health. Higher-status elite institutions are often characterised by having greater per-pupil resources, smaller class sizes and providing well-equipped facilities and wider range of activities (13), which may affect the subsequent health of its attendees through several mechanisms. First, attendance at a high-status institution may lead to improved labour market outcomes (11,14) which in turn may benefit health (15). Net of economic pathways, peers at elite institutions tend to display different patterns in health behaviours and cultural norms (16,17), which could influence the health and health behaviours of those attending these institutions given the importance of peer effects on health behaviours (18–20). Finally, elite institutions may have more cognitively stimulating environment through having smaller class sizes, more experienced teachers and high-achieving peers (21,22); this may benefit cognition in adolescence, across adulthood and ultimately health in midlife (23).

Fewer studies had explicitly addressed this association between type of education and subsequent adult health (24–27). Most studies have found weak evidence of a favourable association between attending a selective school and subsequent health, using self-rated health in mid-adulthood as the main outcome measure (24–27). It was also found that attending an advantageous secondary school measured by indicators such as student-teacher ratio or a highly selective university was associated with better cognitive function in old age (21,22).

Overall, only a few studies had investigated the effect of type of education on subsequent health directly. The classification of the type of education used varied across studies, and only one of the studies had investigated at both school and university levels simultaneously (27). Moreover, self-rated health was used as the measurement for most studies examining overall health, while the association between education type and health differs by health outcome. Also, reporting bias of self-rated health tends to be affected by individual’s education level, implying that the estimated association could be underestimated (28,29). These factors highlight the need for multiple objective outcomes in investigating the association between type of education and health.

Thus, we investigated the association between the type of education and subsequent adult health in mid-adulthood, using data from a UK-wide representative longitudinal study with participants born in 1970 and objective health outcome measures. Multiple health outcomes were investigated, enabling several dimensions of health to be considered and enabling effect sizes to be compared across outcomes (30). Private high school (secondary school) and higher-status university attendance were investigated in particular. We hypothesised that private high school or higher-status university attendance would both be associated with better health in mid-adulthood and this association would only partly be explained by preceding confounders.

## Methods

### Data and sample

We used data from the 1970 British Cohort Study (BCS70), a representative UK cohort born in a single week of 1970 (31). This involved 17 196 babies at baseline, with 10 follow-up sweeps conducted when the cohort members were aged 5, 10, 16, 26, 30, 34, 38, 42, 46 and 51 years. At the tenth sweep (conducted in 2016, when the cohort members were aged between 46 to 48), 12 368 participants were invited to participate and 8 581 (69.4%) were interviewed (32). Trained interviewers made initial contact and conducted core interviews, including a cognitive assessment module. This was followed by nurse visits at which biomeasures such as height and weight, blood pressure and grip strength were collected (32). Informed consent was obtained and ethical approval granted (South East Coast – Brighton and Sussex: 15/LO/1446).

### Measures

Type of high school and university attended: Data on cohort members’ type of high school attended was derived from headteacher’s questionnaire and schools census when the cohort members were aged 16, and a retrospective question asked when cohort member was aged 42 if the former two were missing (33). We categorised school types into private schools, grammar schools, and comprehensive and other schools. Participants with special education needs (N=68) and missing high school types (N=406) were excluded from the sample. For the type of university attended, cohort members who had a degree were asked about their first university attended at age 42 (sweep 9, conducted in 2012) (33). Those who attended their first university from the Russell Group of universities were categorised as attended a higher-status university while those who attended all other universities were categorised as attended a normal-status university. The Russell Group of universities includes 24 self-selected institutions which have often been considered as the most prestigious institutions in the UK, with attendance of these institutions being associated with higher graduate earnings (34). We further classified those who did not attend university into two groups according to their highest qualification obtained (no qualification/GCSEs only; A-Levels/diplomas).

Health outcome measures: Ten health outcomes that capture cardiometabolic risks, physical capabilities, and cognitive function were included in the analysis. All outcomes were measured at the tenth wave of BCS70 when the cohort members were aged between 46 to 48. Body Mass Index (BMI): Cohort members’ height and weight (in centimetres and kilograms, using Leicester stadiometer and Tanita BF-522W scales) were taken during the nurse visit and BMI was later calculated using the objective height and weight (32). Blood pressure: Systolic blood pressure was used, as it has been suggested as a better predictor of cardiovascular risks compared with diastolic blood pressure (35). Three measurements of systolic blood pressure were taken during the nurse visit and the average of the second and third reading was used in the analysis (36). Pulse: Pulse has been used as a common indicator of health, associated with risks of cardiovascular diseases and coronary events (37,38). Three measurements of pulse were taken during the nurse visit and the average of the second and third reading was used in the analysis. Grip strength: Three measurements of grip strength (in kilograms) were taken using a Smedley spring-gauge dynamometer during the nurse visit. The maximum grip strength of the dominant hand was used in the analysis (39). Standing balance: Cohort members were asked to raise one foot off the ground a few inches and balance on their other leg. The number of seconds leg was raised with eyes opened was recorded first and the participants were asked to repeat the task with eyes closed if they reached 30 seconds in the eye-opened task. Cognitive function: Four cognition tests were conducted during the core interview session-word list recall, animal naming, letter cancellation and delayed word list recall. These tests intended to capture attention, memory, verbal fluency, and executive function of the participants (40). The number of words recalled, the number of animals named, or the performance score from each test was recorded and analysed as separate outcomes.

### Potential confounders

Potential confounders collected in BCS70 sweep 3 (at age 10): childhood socioeconomic position (SEP), childhood health and childhood cognitive ability were adjusted, as these factors tend to be associated with both elite institution attendance and subsequent health (33,41,42). Indicators of childhood SEP include parental occupational class (measured by Registrar-General’s Social Classes), parental education level (having degree/not) and household income (categorical gross weekly income). Childhood health was measured by whether the participant has missed school due to illness and presence of a disability that interferes with ordinary life, both measured by binary variable. Childhood cognitive ability was measured by five tests taken at age 10, namely the Shortened Edinburgh Reading Test, Friendly Maths Test, Pictorial Language Comprehension Test (PLCT), Spelling Dictation task and British Ability Scales (BAS), as previously described (40). These tests captured a wide range of cognitive abilities, such as word recognition and general mathematical competence.

### Analytical strategy

First, descriptive statistics were generated, and t-tests and chi2 tests for trend were used to compare health outcomes across different education types. We then used multivariable regression models to examine the association between type of high school or university and each of the health outcomes. To aid comparability across outcomes, z-scores were calculated and used in the analysis. For the regression models, sex was adjusted for given little evidence for sex*education interaction. All other potential confounders were then additionally adjusted for (fully adjusted models). For the analysis of the type of university, high school type was also adjusted for. Full information maximum likelihood (FIML) was used to account for missing data under the assumption of “missing at random” (MAR), which assumes that missingness does not depend on unobserved data given observed data. The use of FIML prevents loss in statistical power and reduces limit bias. To aid comparison across outcomes, the signs of the cardiometabolic outcomes (BMI, blood pressure and pulse) were reversed, hence positive values indicate more favourable outcomes on average.

## Results

### Descriptive statistics

Among the 8107 cohort members, a total of 570 (7.0%) participants attended private school and 308 (3.8%) participants attended grammar school (Table 1). A total of 554 (7.2%) participants attended a higher-status university (Table S1). Participants with more advantageous childhood socioeconomic circumstances (higher parental education, occupation status and household income at age 10), fewer school absence due to illness at age 10 were more likely to attend private schools and higher-status universities. Higher cognitive scores at age 10 is associated with higher attendance of both private and grammar schools (Figure S2).

**Table 1.** Descriptive characteristics of the study sample by high school type

| Characteristics and outcome at 46 years | N | High school attended (N/mean; %/SD) |  |  |
| --- | --- | --- | --- | --- |
|  |  | Comprehensive and other | Grammar | Private |
| Full sample | 8 107 | 7 229 (89.2%) | 308 (3.8%) | 570 (7.0%) |
| Male | 3 892 | 3 430 (88.1%) | 153 (3.9%) | 309 (7.9%)** |
| Female | 4 215 | 3 799 (90.1%) | 155 (3.7%) | 261 (6.2%) |
| **Potential confounders** |  |  |  |  |
| Parental education: degree (10y) | 6 602 |  |  |  |
| Yes | 1 146 | 852 (74.4%) | 86 (7.5%) | 208 (18.2%)** |
| No | 5 456 | 5 084 (93.2%) | 159 (2.9%) | 213 (3.9%) |
| Parental occupational class (10y) | 6 922 |  |  |  |
| I Professional | 467 | 319 (68.3%) | 34 (7.3%) | 114 (24.4%)** |
| II Managerial and technical | 1 851 | 1 540 (83.2%) | 104 (5.6%) | 207 (11.2%) |
| III Non manual | 799 | 707 (88.5%) | 41 (5.1%) | 51 (6.4%) |
| III Manual | 2 732 | 2 622 (96.0%) | 57 (2.1%) | 53 (2.0%) |
| IV Partly-skilled | 848 | 813 (96.0%) | 21 (2.4%) | 14 (1.6%) |
| V Unskilled | 225 | 221 (98.2%) | 3 (1.3%) | 1 (0.4%) |
| Weekly household income (10y) | 6 569 |  |  |  |
| Under £35 | 95 | 90 (94.7%) | 2 (2.1%) | 3 (3.2%)** |
| £35-49 | 272 | 255 (93.8%) | 13 (4.8%) | 4 (1.5%) |
| £50-99 | 1 794 | 1 718 ((95.8%) | 44 (2.5%) | 32 (1.8%) |
| £100-149 | 2 317 | 2 183 (94.2%) | 69 (3.0%) | 65 (2.8%) |
| £150-199 | 1 160 | 1 048 (90.3%) | 41 (3.5%) | 71 (6.1%) |
| £200-249 | 478 | 372 (77.8%) | 37 (7.7%) | 69 (14.5%) |
| £250 | 453 | 265 (58.5%) | 35 (7.7%) | 153 (33.8%) |
| Reading score (10y) | 6 042 | 41.7 (11.7) | 51.3 (8.7) | 50.8 (10.0)** |
| Maths score (10y) | 6 041 | 45.3 (11.3) | 55.0 (9.2) | 54.5 (10.8)** |
| Spelling Dictation task (10y) | 6 464 | 35.7 (10.1) | 42.2 (7.1) | 41.2 (8.0)** |
| Pictorial Language Comprehension (10y) | 6 575 | 62.1 (10.11) | 69.8 (9.5) | 69.1 (10.5)** |
| British Ability Scales (10y) | 6 010 | 61.1 (12.5) | 72.2 (11.1) | 71.4 (12.6)** |
| School missed due to illness (10y) | 7 063 |  |  |  |
| Yes | 2 611 | 2 400 (91.9%) | 84 (3.2%) | 127 (4.9%)** |
| No | 4 452 | 3 952 (88.8%) | 180 (4.0%) | 320 (7.2%) |
| Presence of disability (10y) | 7 077 |  |  |  |
| Yes | 453 | 409 (90.2%) | 12 (2.7%) | 32 (7.1%) |
| No | 6 624 | 5 958 (90.0%) | 249 (3.8%) | 417 (6.3%) |
| **Outcomes** |  |  |  |  |
| Cardiometabolic |  |  |  |  |
| Body mass index (kg/m<sup>2</sup>) | 7 068 | 28.6 (5.5) | 27.7 (5.4) | 26.9 (4.8)** |
| Systolic blood pressure (mmHg) | 7 180 | 124.2 (15.2) | 123.6 (14.2) | 122.0 (14.3)** |
| Pulse (beats per minute) | 7 178 | 68.4 (10.6) | 66.7 (10.5) | 66.3 (10.3)** |
| Physical function |  |  |  |  |
| Grip strength (kg) | 7 146 | 38.0 (12.1) | 38.6 (10.9) | 39.5 (11.1)** |
| Standing balance: eyes opened (s) | 7 054 | 28.1 (5.8) | 28.4 (5.6) | 28.8 (4.9)* |
| Standing balance: eyes closed (s) | 6 118 | 11.6 (9.6) | 14.2 (10.4) | 14.3 (10.3)** |
| Cognitive function |  |  |  |  |
| Immediate word recall | 8 052 | 6.6 (1.4) | 7.1 (1.4) | 7.2 (1.3)** |
| Animal naming | 8 047 | 23.5 (6.0) | 26.0 (6.8) | 26.2 (6.4)** |
| Letter cancellation speed | 7 822 | 345.5 (84.1) | 351.6 (76.7) | 361.6 (82.7)** |
| Delayed word recall | 8 046 | 5.4 (1.8) | 6.2 (1.7) | 6.1 (1.8)** |

\*P<0.05, \*\*P<0.01, using chi-square or t-test across high school type

### High school type and health outcomes

Overall, private school attendance was associated with better health outcomes in most cardiometabolic and cognitive outcomes compared with comprehensive school attendance in sex-adjusted models (Figure 1/Table S2). In sex-adjusted models, private school attendance had 0.32 (95%CI: 0.23, 0.41) and 0.19 (0.10, 0.27) better outcome (lower z-scores) in BMI and systolic blood pressure. After full adjustment, these estimates have attenuated about 50% (BMI: 0.14, 0.04, 0.23; systolic blood pressure: 0.10, 0.01, 0.19). Private school attendees also performed better in the letter cancellation task (0.11, 0.02, 0.20) after fully adjusted.

**Figure 1.**
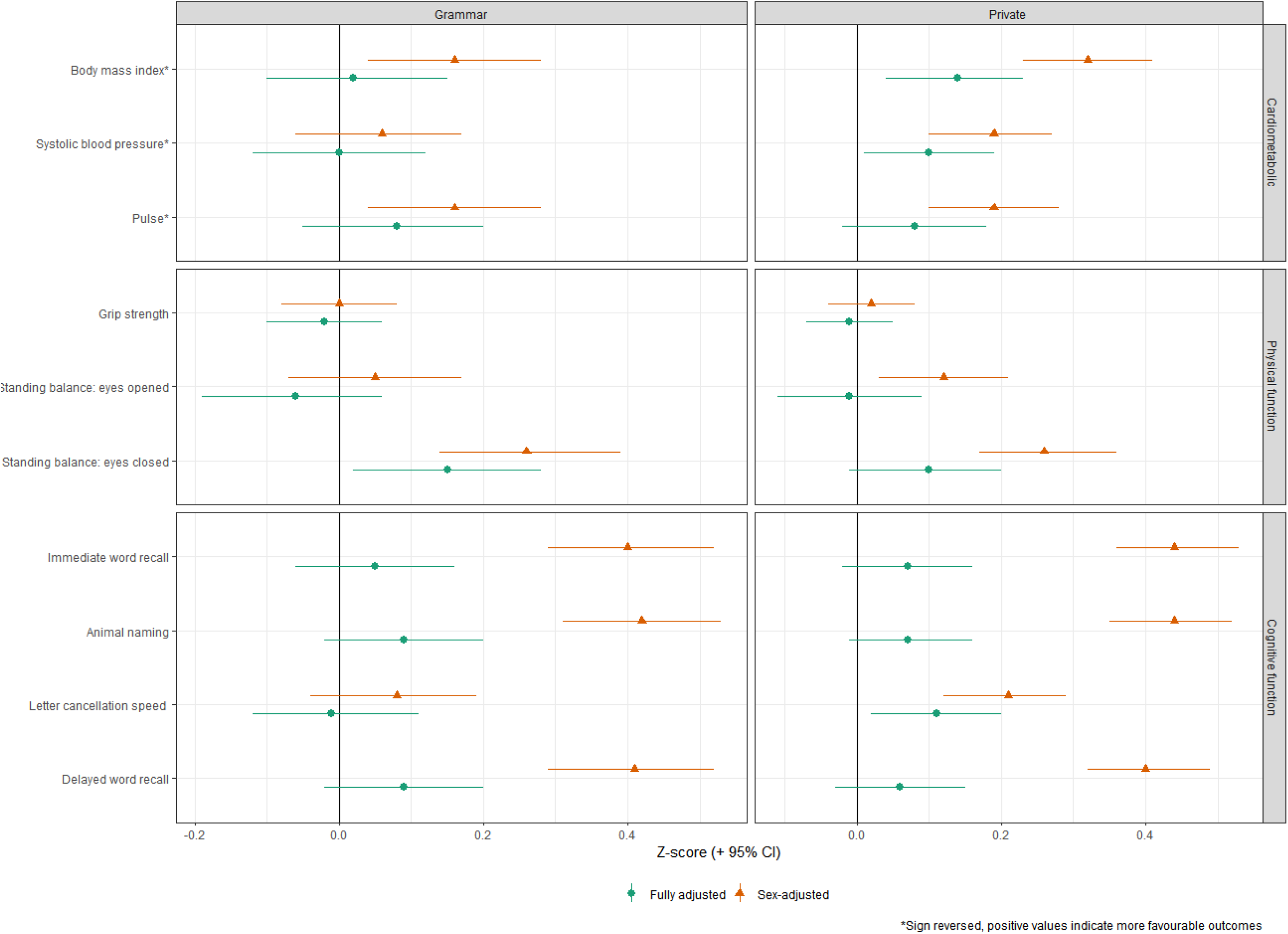
Associations between type of high school attended and health outcomes at age 46, comprehensive/other as reference group

Grammar school attendance was also associated with better outcomes in BMI, pulse, eye-closed standing balance, and most of the cognitive tasks compared with comprehensive school attendance in sex-adjusted models. However, most of these associations attenuated after fully adjustment, except for eye-closed standing balance (0.15, 0.02, 0.28). No major difference was found between grammar school and private school attendance, except for BMI in sex-adjusted model, which private school attendance being associated with better outcome in BMI (0.16, 0.01, 0.31) (Table S3).

### Qualification/university type and health outcomes

In sex-adjusted models, higher-status university attendance was associated with better outcomes/performance in BMI and most cognitive task (except for letter cancellation) compared with normal-status university attendance (Figure 2/Table S4). After adjustment, the association with BMI attenuated, while the associations with the three cognitive tasks remained (Immediate word recall: 0.16, 0.07, 0.26; Animal naming: 0.10, 0.00, 0.19; Delayed word recall: 0.14, 0.05, 0.24).

**Figure 2.**
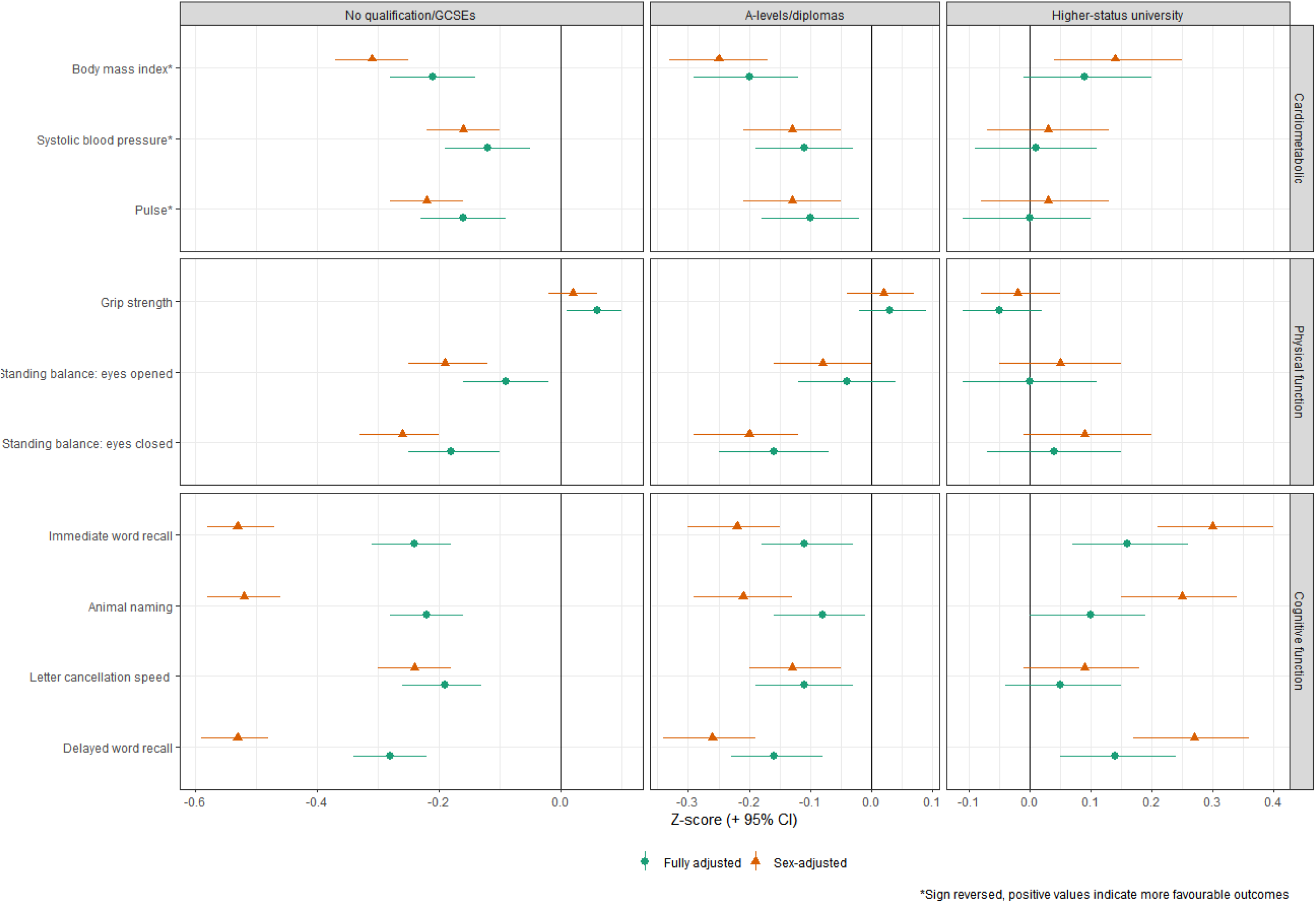
Associations between type of qualification/university attended and health outcomes at age 46, normal status university as reference group

In sex-adjusted models, having no degree was associated with worse outcomes in most health measures compared with normal-status university attendance, except for grip strength (for both with no qualifications/GCSEs only and with A-levels/diplomas) and eyes-opened standing balance (A-levels/diplomas only). These associations remained after being fully adjusted for potential confounders.

## Discussion

### Main findings

The findings indicate a favourable link between attending elite educational institutions and health at age 46 in the BCS70 cohort. Private school attendance was associated with better cardiometabolic outcomes than comprehensive school attendance, after accounting for various potential confounders. Attending higher-status universities was associated with better cognitive function, while having no degree was linked to poorer health compared with normal-status university attendance. These results emphasise the importance of considering other aspects of education when examining its effects on health.

### Comparison with previous findings

Previous British studies using the 1958 National Child Development Study (NCDS) or the Aberdeen Children of the 1950s birth cohort study have found a positive but insignificant association between attending a selective school and self-rated health in later life (24–26). These studies utilised the comprehensive reform in the UK as natural experiment, comparing those exposed to a selective or non-selective schooling system, which is equivalent to the distinction between grammar schools and comprehensive schools. An analysis of our data suggested that there was not a clear discontinuity which could be used in such a design (see Figure S2). Nevertheless, our findings also suggest that many beneficial associations between grammar school attendance and health have attenuated after fully adjustment consistent with these studies. A previous study using the BCS70 cohort concluded that private school and high-status university attendance was associated with lower self-reported BMI (27). Our analyses have extended this previous study by using objective health outcome measures and also found consistent evidence for BMI.

Previous studies in the US have found that attending an advantageous secondary school measured by indicators such as student-teacher ratio or a highly selective university was associated with better cognitive function in old age (21,22). This is consistent with our finding that higher-status university attendance was associated with better performance in most cognitive tasks. However, these studies have used different measures of education type and cohorts, and have focused on cognitive function in old age, making the results less comparable.

### Explanation of findings

Several potential mechanisms may explain the beneficial associations between elite education attendance and subsequent health. At the high school level, attending private schools was associated with lower BMI and systolic blood pressure at age 46. Private schools can promote healthier health-related behaviours among their attendees. Peers at private schools tend to display different health behaviours such as greater physical activity (16,17), which can influence attendees’ behaviours through peer effect (18,20). Such behaviours tend to persist into adulthood (43,44), which contributes to the beneficial associations found between private school attendance and health. Apart from behaviour factors, private school attendance is often associated with higher subsequent socioeconomic positions (11,14). This can favourably associate with BMI and blood pressure, either directly by increasing the affordability of healthy diet/activity or indirectly through better social connections and neighbourhood environment (45). Finally, our results could be explained by confounding; while we accounted for multiple potential prospectively ascertained confounders, the available measures may have imperfectly captured the relevant constructs (e.g., childhood cognition or parental SEP), thus leading to residual confounding. Conversely, however, inclusion of measures of childhood cognitive function may have induced an overadjustment bias as cognitive function is likely to be related to educational resources and quality even within the categories defined by our exposure variables – e.g. within private schools or Russell Group universities.

At the university level, higher-status university attendance was found to be associated with better cognitive function after full adjustment; associations with other outcomes were largely attenuated after adjustment. This contrasts with findings for the lowest education group, in which associations with outcomes remained after adjustment. The distinction in factors associated with health such as subsequent socioeconomic position may be greater between those with and without a degree than between those who attended different university types. In our study sample, 7.2% of the participants attended a high-status university; while this is a similar fraction of those who attended private schools, the composition of the group likely differs in ways which linked with subsequent health. The persisting links with cognition outcomes likely partly reflects the selection into higher status institutions (confounding due to cognition). It could also partly reflect causal effects on cognition (46), due for example to the enhanced pathways of cognitively stimulating environments that higher subsequent SEP affords (eg, higher status occupations enabled by higher status institutions attended). Comparing the subject of degrees attended may offer some insight into this in future, since the economic returns to higher education markedly differ by degree attended (47).

### Strengths and limitations

The strengths of this study include the usage of a large, nationally-representative study cohort with detailed information on the type of high school and university attended, multiple health outcomes in middle age and a range of childhood socioeconomic, health and cognitive measures. The health outcomes investigated were measured objectively during a nurse visit (32). This reduces potential reporting bias using self-reported health measures, as health reporting tends to differ according to education attainment (48,49). Moreover, multiple health outcomes were investigated simultaneously. This enables different aspects of health to be investigated, which enables the education-health association to be better understood given its potentially heterogeneous nature.

Our study has several limitations. During the nurse visits, cohort members were first invited to perform the eye-opened standing balance and only those who reached 30 seconds were further invited to perform the eye-closed standing balance (32). Therefore, results of eye-closed standing balance were based on a more selective group which might explain the different results found between the two standing balance tasks. Apart from the outcome measures, results from this study are subjected to potential confounding due to the existence of unmeasured factors that can affect participants’ type of school/university attended and subsequent health outcomes. For example, potential confounders such as childhood SEP and cognitive function are difficult to be captured comprehensively and factors such as parents’ level of motivation regarding education were unobserved. Therefore, residual confounding may exist, and the causal nature of the relationship is difficult to establish. Future studies could use other designs such as quasi-experimental approaches, where exogenous variation in elite education access can be identified.

Finally, this study investigated the effects of higher-status education attendance in one particular generation in the UK. With the changing school fees, economic returns and motivation of choosing elite education (12,50,51), the results may differ in other cohorts and contexts. For example, a significant rise in both school fees and labour market return in the UK was observed in recent decades (12,51), implying that its beneficial associations with health may become stronger in more recently born cohorts.

### Potential implications

Our findings suggest that type of education may play an important part in the education-health association. Previous studies investigating the effects of education on health have predominantly focused on educational attainment (1,2,7–9). As the type of education was uncaptured, the effects of education are likely to be underestimated. Moreover, if this association is causal, future policies aimed at reducing health inequalities could take education quality into account as well as attainment. This is particularly important given the increases in university attendance, in which other aspects of the education experience may better distinguish health inequality.

## Conclusion

Results from the current study suggest a beneficial association between higher-status institution attendance and subsequent health in mid-adulthood, using multiple objective health measures. The type of education institution attended may play an important role in health inequalities. Further research is warranted to test the causal nature of this relationship and its generalisability to more recently born cohorts given changes in secondary education and the expansion of higher education.

## Supporting information

(all supplementary materials included)

## Data Availability

The datasets analysed in this study are available from the UK Data Service website:https://ukdataservice.ac.uk/

## References

1. Von dem Knesebeck O, Verde PE, Dragano N. Education and health in 22 European countries. Social science & medicine. 2006 Sep 1;63(5):1344–51.

2. Montez JK, Friedman EM. Educational attainment and adult health: Under what conditions is the association causal? Social Science & Medicine. 2015 Feb 1;127:1–7.

3. Conti G, Heckman J, Urzua S. The education-health gradient. American Economic Review. 2010 May;100(2):234–38.

4. Davies NM, Dickson M, Smith GD, van den Berg G, Windmeijer F. The causal effects of education on health, mortality, cognition, well-being, and income in the UK Biobank. bioRxiv. 2016 Jan 1:074815.

5. Mouw T, Koster A, Wright ME, Blank MM, Moore SC, Hollenbeck A, Schatzkin A. Education and risk of cancer in a large cohort of men and women in the United States. PloS one. 2008 Nov 4;3(11):e3639.

6. Pathirana TI, Jackson CA. Socioeconomic status and multimorbidity: a systematic review and meta-analysis. Australian and New Zealand journal of public health. 2018 Apr;42(2):186–94.

7. Braakmann N. The causal relationship between education, health and health related behaviour: Evidence from a natural experiment in England. Journal of Health Economics. 2011 Jul 1;30(4):753–63.

8. Silles MA. The causal effect of education on health: Evidence from the United Kingdom. Economics of Education Review. 2009 Feb 1;28(1):122–8.

9. Howe L, Rasheed H, Jones PR, Boomsma DI, Evans DM, Giannelis A, Hayward C, Hopper JL, Hughes A, Lahtinen H, Li S. Educational attainment, health outcomes and mortality: a within-sibship Mendelian randomization study. medRxiv. 2022 Jan 1.

10. Ryan C, Sibieta L. Private Schooling in the UK and Australia. 2010.

11. Green F, Henseke G, Vignoles A. Private schooling and labour market outcomes. British Educational Research Journal. 2017 Feb 1;43(1):7–28.

12. Green F, Anders J, Henderson M, Henseke G. Who Chooses Private Schooling in Britain and Why? 2017.

13. Anders J, Green F, Henderson M, Henseke G. Determinants of private school participation: All about the money?. British Educational Research Journal. 2020 Oct;46(5):967–92.

14. Hoekstra M. The Effect of Attending the Flagship State University on Earnings: A Discontinuity-Based Approach. The Review of Economics and Statistics. 2009 Nov 1;91(4):717–24.

15. Arber S, Fenn K, Meadows R. Subjective financial well-being, income and health inequalities in mid and later life in Britain. Social Science & Medicine. 2014 Jan 1;100:12–20.

16. Butikofer A, Ginja R, Landaud F, Løken KV. School selectivity, peers, and mental health. NHH Dept. of Economics Discussion Paper. 2020 Oct 14(21).

17. Walsemann KM, Gee GC, Ro A. Educational Attainment in the Context of Social Inequality: New Directions for Research on Education and Health. American Behavioral Scientist. 2013;57(8):1082–104.

18. Beal AC, Ausiello J, Perrin JM. Social influences on health-risk behaviors among minority middle school students. Journal of Adolescent Health. 2001 Jun 1;28(6):474–80.

19. Fletcher JM. Peer influences on adolescent alcohol consumption: evidence using an instrumental variables/fixed effect approach. Journal of Population Economics. 2012 Oct;25(4):1265–86.

20. Kirby J, Levin KA, Inchley J. Parental and peer influences on physical activity among Scottish adolescents: a longitudinal study. Journal of physical activity & health. 2011 Aug 1;8(6).

21. Garcia S, Moorman SM. College Selectivity and Later-Life Memory Function: Evidence From the Wisconsin Longitudinal Study. Research on Aging. 2021 Jan 1;43(1):14–24.

22. Moorman SM, Greenfield EA, Garcia S. School Context in Adolescence and Cognitive Functioning 50 Years Later. Journal of Health and Social Behavior. 2019 Dec 1;60(4):493–508.

23. Gow AJ, Johnson W, Pattie A, Brett CE, Roberts B, Starr JM, Deary IJ. Stability and change in intelligence from age 11 to ages 70, 79, and 87: the Lothian Birth Cohorts of 1921 and 1936. Psychology and aging. 2011 Mar;26(1):232.

24. Basu A, Jones AM, Dias PR. Heterogeneity in the impact of type of schooling on adult health and lifestyle. Journal of Health Economics. 2018 Jan 1;57:1–14.

25. Butler J, Black C, Craig P, Dibben C, Dundas R, Hilton Boon M, Johnston M, Popham F. The long-term health effects of attending a selective school: a natural experiment. BMC medicine. 2020 Dec;18(1):1–15.

26. Jones AM, Pastore C, Rice N. Tracking pupils into adulthood: selective schools and long-term well-being in the 1958 British cohort. HEDG, c/o Department of Economics, University of York; 2018 Nov.

27. Bann D, Hamer M, Parsons S, Ploubidis GB, Sullivan A. Does an elite education benefit health? Findings from the 1970 British Cohort Study. International Journal of Epidemiology. 2017 Feb 1;46(1):293–302.

28. Bago d’Uva T, O’Donnell O, van doorslaer E. Differential health reporting by education level and its impact on the measurement of health inequalities among older Europeans. International Journal of Epidemiology. 2008 Dec 1;37(6):1375–83.

29. Dowd JB, Todd M. Does Self-reported Health Bias the Measurement of Health Inequalities in U.S. Adults? Evidence Using Anchoring Vignettes From the Health and Retirement Study. The Journals of Gerontology: Series B. 2011 Jul 1;66B(4):478–89.

30. VanderWeele TJ, Mathur MB, Chen Y. Outcome-Wide Longitudinal Designs for Causal Inference: A New Template for Empirical Studies. 2020 Aug 1;35(3):437–66.

31. Sullivan A, Brown M, Hamer M, Ploubidis GB. Cohort Profile Update: The 1970 British Cohort Study (BCS70). International journal of epidemiology.:dyac148.

32. Brown M, Peters A. 1970 British Cohort Study Age 46 Survey User Guide. Centre for Longitudinal Studies, UCL, London.2019.

33. Sullivan A, Parsons S, Wiggins R, Heath A, Green F. Social origins, school type and higher education destinations. Oxford Review of Education. 2014 Nov 2;40(6):739–63.

34. Chevalier A, Conlon G. Does it pay to attend a prestigious university?. Available at SSRN 435300. 2003 Aug.

35. Strandberg TE, Pitkala K. What is the most important component of blood pressure: systolic, diastolic or pulse pressure?. Current opinion in nephrology and hypertension. 2003 May 1;12(3):293–7.

36. Hackam DG, Quinn RR, Ravani P, Rabi DM, Dasgupta K, Daskalopoulou SS, et al. The 2013 Canadian Hypertension Education Program Recommendations for Blood Pressure Measurement, Diagnosis, Assessment of Risk, Prevention, and Treatment of Hypertension. Canadian Journal of Cardiology. 2013 May 1;29(5):528–42.

37. Kannel WB, Kannel C, Paffenbarger RS, Cupples LA. Heart rate and cardiovascular mortality: The Framingham study. Am Heart J. 1987 Jun 1;113(6):1489–94.

38. Jensen MT. Resting heart rate and relation to disease and longevity: past, present and future. Scandinavian journal of clinical and laboratory investigation. 2019 Feb 17;79(1-2):108–16.

39. Roberts HC, Denison HJ, Martin HJ, Patel HP, Syddall H, Cooper C, et al. A review of the measurement of grip strength in clinical and epidemiological studies: towards a standardised approach. 2011 Jul 1;40(4):423–9.

40. Cognitive measures in the 1970 British Cohort Study - CLOSER [Internet]. [cited 2023 May 4]. Available from: https://closer.ac.uk/cross-study-data-guides/cognitive-measures-guide/bcs70-cognition/

41. Cohen S, Janicki-Deverts D, Chen E, Matthews KA. Childhood socioeconomic status and adult health. Ann N Y Acad Sci. 2010 Feb 1;1186(1):37–55.

42. Delaney L, Smith JP. Childhood health: trends and consequences over the life-course. The Future of Children/Center for the Future of Children, the David and Lucile Packard Foundation. 2012;22(1):43.

43. Craigie AM, Lake AA, Kelly SA, Adamson AJ, Mathers JC. Tracking of obesity-related behaviours from childhood to adulthood: A systematic review. Maturitas. 2011 Nov;70(3):266–84.

44. Tammelin R, Yang X, Leskinen E, Kankaanpaa A, Hirvensalo M, Tammelin T, Raitakari OT. Tracking of physical activity from early childhood through youth into adulthood. Med Sci Sports Exerc. 2014;46(5):955–62.

45. Adler NE, Newman K. Socioeconomic disparities in health: pathways and policies. Health affairs. 2002 Mar;21(2):60–76.

46. Ritchie SJ, Tucker-Drob EM. How much does education improve intelligence? A meta-analysis. Psychological science. 2018 Aug;29(8):1358–69.

47. Britton J, Dearden L, Shephard N, Vignoles A. How English domiciled graduate earnings vary with gender, institution attended, subject and socio-economic background. IFS working papers; 2016.

48. Bago d’Uva T, O’Donnell O, van doorslaer E. Differential health reporting by education level and its impact on the measurement of health inequalities among older Europeans. Int J Epidemiol. 2008 Dec;37(6):1375–83.

49. Bago d’Uva T, Lindeboom M, O’Donnell O, van Doorslaer E. Education-related inequity in healthcare with heterogeneous reporting of health. J R Stat Soc Ser A Stat Soc. 2011 Jul;174(3):639–64.

50. Burgess S, Greaves E, Vignoles A. School choice in England: evidence from national administrative data. Oxford Review of Education. 2019 Sep 3;45(5):690–710.

51. Green F, Machin S, Murphy R, Zhu Y. The changing economic advantage from private schools. Economica. 2012 Oct;79(316):658–79.

