## Supplementary material for "Associations of schooling type, qualification type, and subsequent health in mid-adulthood: Evidence from the 1970 British Cohort Study": (all supplementary materials included)

Table S1. Descriptive characteristics of the study sample by qualification/university type

| Characteristics and outcome at 46 years | N | No qualification/GCSEs | A-levels/diplomas | Normal-status university | High-status university |
|---|---|---|---|---|---|
| Full sample | 7 685 | 4 616 (60.1%) | 1 134 (14.7%) | 1 381 (17.8%) | 554 (7.2%) |
| Male | 3 686 | 2 297 (62.3%) | 475 (12.9%) | 642 (17.4%) | 272 (7.4%)** |
| Female | 3 999 | 2 319 (58.0%) | 659 (16.5%) | 739 (18.5%) | 282 (7.1%) |
| Parental education: degree (10y) | 6 249 | | | | |
| Yes | 1 093 | 370 (33.9%) | 162 (14.8%) | 352 (32.2%) | 209 (19.1%)** |
| No | 5 156 | 3 394 (65.8%) | 769 (14.9%) | 747 (14.5%) | 246 (4.8%) |
| Parental occupational class (10y) | 6 544 | | | | |
| I Professional | 435 | 115 (26.4%) | 71 (16.3%) | 157 (36.1%) | 92 (21.2%)** |
| II Managerial and technical | 1 746 | 807 (46.2%) | 282 (16.2%) | 439 (25.2%) | 218 (12.5%) |
| III Non manual | 758 | 424 (55.9%) | 122 (16.1%) | 155 (20.5%) | 57 (7.5%) |
| III Manual | 2 570 | 1 799 (70.0%) | 353 (13.7%) | 321 (12.5%) | 97 (3.8%) |
| IV Partly-skilled | 816 | 579 (71.0%) | 122 (15.0%) | 94 (11.5%) | 21 (2.6%) |
| V Unskilled | 219 | 174 (79.5%) | 25 (11.4%) | 17 (7.8%) | 3 (1.4%) |
| Weekly household income (10y) | 6 203 | | | | |
| Under £35 | 92 | 65 (70.7%) | 10 (10.9%) | 15 (16.3%) | 2 (2.2%)** |
| £35-49 | 265 | 197 (74.3%) | 35 (13.2%) | 29 (10.9%) | 4 (1.5%) |
| £50-99 | 1 677 | 1 171 (69.8%) | 217 (12.9%) | 236 (14.1%) | 53 (3.2%) |
| £100-149 | 2 186 | 1 389 (63.4%) | 332 (15.2%) | 341 (15.6%) | 124 (5.7%) |
| £150-199 | 1 108 | 582 (52.5%) | 181 (16.3%) | 238 (21.5%) | 107 (9.7%) |
| £200-249 | 449 | 173 (38.5%) | 87 (19.4%) | 118 (26.3%) | 71 (15.8%) |
| £250 | 426 | 147 (34.5%) | 57 (13.4%) | 119 (27.9%) | 103 (24.2%) |
| Reading score (10y) | 5 702 | 39.0 (11.6) | 44.6 (10.6) | 48.8 (9.8) | 52.8 (8.5)** |
| Maths score (10y) | 5 707 | 42.7 (11.0) | 47.5 (10.5) | 52.5 (9.8) | 56.8 (9.1)** |
| Spelling Dictation task (10y) | 6 092 | 34.1 (10.5) | 37.6 (9.4) | 39.8 (8.2) | 42.4 (7.5)** |
| Pictorial Language Comprehension (10y) | 6 202 | 60.3 (9.8) | 64.1 (10.1) | 67.1 (9.8) | 70.4 (9.6)** |
| British Ability Scales (10y) | 5 674 | 58.3 (12.0) | 64.0 (11.8) | 68.4 (11.7) | 73.7 (10.9)** |
| School missed due to illness (10y) | 6 677 | | | | |
| Yes | 2 464 | 1 541 (62.5%) | 372 (15.1%) | 403 (16.4%) | 148 (6.0%)** |
| No | 4 213 | 2 447 (58.1%) | 619 (14.7%) | 800 (19.0%) | 347 (8.2%) |
| Presence of disability (10y) | 6 690 | | | | |
| Yes | 452 | 301 (66.6%) | 59 (13.1%) | 70 (15.5%) | 22 (4.9%)** |
| No | 6 238 | 3 703 (59.4%) | 932 (14.9%) | 1 128 (18.1%) | 475 (7.6%) |
| Body mass index (kg/$m^2$) | 6 715 | 28.9 (5.6) | 28.6 (5.5) | 27.3 (4.9) | 26.5 (4.7)** |
| Systolic blood pressure (mmHg) | 6 829 | 124.9 (15.2) | 123.7 (15.3) | 122.1 (14.3) | 121.9 (14.6)** |
| Pulse (beats per minute) | 6 827 | 69.0 (10.8) | 68.1 (10.1) | 66.7 (10.2) | 66.3 (11.0)** |
| Grip strength (kg) | 6 791 | 38.6 (12.5) | 37.1 (11.6) | 37.6 (11.0) | 38.1 (10.8)** |
| Standing balance: eyes opened (seconds) | 6 698 | 27.7 (6.3) | 28.3 (5.5) | 28.8 (4.7) | 29.1 (4.2)** |
| Standing balance: eyes closed (seconds) | 5 805 | 11.2 (9.3) | 11.6 (9.4) | 13.7 (10.5) | 14.6 (10.3)** |
| Immediate word recall (number) | 7 630 | 6.3 (1.4) | 6.8 (1.4) | 7.1 (1.4) | 7.5 (1.3)** |
| Animal naming (number) | 7 625 | 22.5 (5.8) | 24.4 (6.0) | 25.7 (6.3) | 27.2 (6.0)** |
| Letter cancellation speed | 7 412 | 339.1 (84.3) | 350.2 (80.1) | 360.0 (87.3) | 366.5 (82.7)** |
| Delayed word recall (number) | 7 623 | 5.1 (1.7) | 5.7 (1.7) | 6.1 (1.7) | 6.6 (1.7)** |

*P<0.05, **P<0.01, using chi-square or t-test across university type

Table S2. Associations between type of high school attended and health outcomes at age 46, comprehensive and other as reference group.

| Outcomes at 46 years | N | Grammar; Z-score (95% CI) | | Private; Z-score (95% CI) | |
|---|---|---|---|---|---|
| | | Sex-adjusted models | Fully adjusted models | Sex-adjusted models | Fully adjusted models |
| **-Cardiometabolic** | | | | | |
| Body mass index | 7 068 | 0.16 (0.04, 0.28) | 0.02 (-0.10, 0.15) | 0.32 (0.23, 0.41) | 0.14 (0.04, 0.23) |
| Systolic blood pressure | 7 180 | 0.06 (-0.06, 0.17) | -0.00 (-0.12, 0.12) | 0.19 (0.10, 0.27) | 0.10 (0.01, 0.19) |
| Pulse | 7 178 | 0.16 (0.04, 0.28) | 0.08 (-0.05, 0.20) | 0.19 (0.10, 0.28) | 0.08 (-0.02, 0.18) |
| **-Physical function** | | | | | |
| Grip strength | 7 146 | 0.00 (-0.08, 0.08) | -0.02 (-0.10, 0.06) | 0.02 (-0.04, 0.08) | -0.01 (-0.07, 0.05) |
| Standing balance: eyes opened | 7 054 | 0.05 (-0.07, 0.17) | -0.06 (-0.19, 0.06) | 0.12 (0.03, 0.21) | -0.01 (-0.11, 0.09) |
| Standing balance: eyes closed | 6 118 | 0.26 (0.14, 0.39) | 0.15 (0.02, 0.28) | 0.26 (0.17, 0.36) | 0.10 (-0.01, 0.20) |
| **-Cognitive function** | | | | | |
| Immediate word recall | 8 052 | 0.40 (0.29, 0.52) | 0.05 (-0.06, 0.16) | 0.44 (0.36, 0.53) | 0.07 (-0.02, 0.16) |
| Animal naming | 8 047 | 0.42 (0.31, 0.53) | 0.09 (-0.02, 0.20) | 0.44 (0.35, 0.52) | 0.07 (-0.01, 0.16) |
| Letter cancellation speed | 7 822 | 0.08 (-0.04, 0.19) | -0.01 (-0.12, 0.11) | 0.21 (0.12, 0.29) | 0.11 (0.02, 0.20) |
| Delayed word recall | 8 046 | 0.41 (0.29, 0.52) | 0.09 (-0.02, 0.20) | 0.40 (0.32, 0.49) | 0.06 (-0.03, 0.15) |

Table S3. Associations between type of high school attended and health outcomes at age 46, grammar school as reference group

| Outcomes at 46 years | N | Comprehensive and other<br>Z-score (95% CI) | | Private<br>Z-score (95% CI) | |
|---|---|---|---|---|---|
| | | Sex-adjusted models | Fully adjusted models | Sex-adjusted models | Fully adjusted models |
| **-Cardiometabolic** | | | | | |
| Body mass index | 7 068 | -0.16 (-0.28, -0.04) | -0.02 (-0.14, 0.10) | 0.16 (0.01, 0.31) | 0.12 (-0.03, 0.26) |
| Systolic blood pressure | 7 180 | -0.06 (-0.17, 0.06) | 0.00 (-0.12, 0.12) | 0.13 (-0.01, 0.27) | 0.10 (-0.04, 0.24) |
| Pulse | 7 178 | -0.16 (-0.28, -0.04) | -0.08 (-0.20, 0.05) | 0.03 (-0.11, 0.18) | 0.00 (-0.14, 0.15) |
| **-Physical function** | | | | | |
| Grip strength | 7 146 | 0.00 (-0.08, 0.08) | 0.02 (-0.06, 0.10) | 0.02 (-0.07, 0.12) | 0.01 (-0.09, 0.10) |
| Standing balance: eyes opened | 7 054 | -0.05 (-0.17, 0.07) | 0.06 (-0.06, 0.19) | 0.07 (-0.08, 0.22) | 0.05 (-0.09, 0.20) |
| Standing balance: eyes closed | 6 118 | -0.26 (-0.39, -0.14) | -0.15 (-0.28, -0.02) | -0.00 (-0.16, 0.15) | -0.05 (-0.21, 0.10) |
| **-Cognitive function** | | | | | |
| Immediate word recall | 8 052 | -0.40 (-0.52, -0.29) | -0.05 (-0.16, 0.06) | 0.04 (-0.10, 0.18) | 0.02 (-0.11, 0.15) |
| Animal naming | 8 047 | -0.42 (-0.53, -0.31) | -0.09 (-0.20, 0.02) | 0.02 (-0.12, 0.16) | -0.01 (-0.14, 0.12) |
| Letter cancellation speed | 7 822 | -0.08 (-0.19, 0.04) | 0.01 (-0.11, 0.12) | 0.13 (-0.01, 0.27) | 0.11 (-0.03, 0.25) |
| Delayed word recall | 8 046 | -0.41 (-0.52, -0.29) | -0.09 (-0.20, 0.02) | -0.01 (-0.14, 0.13) | -0.03 (-0.16, 0.10) |

Table S4. Associations between type of qualification/university attended and health outcomes at age 46, normal-status university as reference group

| Outcomes at 46 years | N | No qualification/GCSEs Z-score (95%CI) | | A-levels/diplomas Z-score (95%CI) | | Higher-status university Z-score (95%CI) | |
|---|---|---|---|---|---|---|---|
| | | Sex-adjusted models | Fully adjusted models | Sex-adjusted models | Fully adjusted models | Sex-adjusted models | Fully adjusted models |
| **-Cardiometabolic** | | | | | | | |
| Body mass index | 6 715 | -0.31 (-0.37, -0.25) | -0.21 (-0.28, -0.14) | -0.25 (-0.33, -0.17) | -0.20 (-0.29, -0.12) | 0.14 (0.04, 0.25) | 0.09 (-0.01, 0.20) |
| Systolic blood pressure | 6 829 | -0.16 (-0.22, -0.10) | -0.12 (-0.19, -0.05) | -0.13 (-0.21, -0.05) | -0.11 (-0.19, -0.03) | 0.03 (-0.07, 0.13) | 0.01 (-0.09, 0.11) |
| Pulse | 6 827 | -0.22 (-0.28, -0.16) | -0.16 (-0.23, -0.09) | -0.13 (-0.21, -0.05) | -0.10 (-0.18, -0.02) | 0.03 (-0.08, 0.13) | -0.00 (-0.11, 0.10) |
| **-Physical function** | | | | | | | |
| Grip strength | 6 791 | 0.02 (-0.02, 0.06) | 0.06 (0.01, 0.10) | 0.02 (-0.04, 0.07) | 0.03 (-0.02, 0.09) | -0.02 (-0.08, 0.05) | -0.05 (-0.11, 0.02) |
| Standing balance: eyes opened | 6 698 | -0.19 (-0.25, -0.12) | -0.09 (-0.16, -0.02) | -0.08 (-0.16, 0.00) | -0.04 (-0.12, 0.04) | 0.05 (-0.05, 0.15) | -0.00 (-0.11, 0.11) |
| Standing balance: eyes closed | 5 805 | -0.26 (-0.33, -0.20) | -0.18 (-0.25, -0.10) | -0.20 (-0.29, -0.12) | -0.16 (-0.25, -0.07) | 0.09 (-0.01, 0.20) | 0.04 (-0.07, 0.15) |
| **-Cognitive function** | | | | | | | |
| Immediate word recall | 7 630 | -0.53 (-0.58, -0.47) | -0.24 (-0.31, -0.18) | -0.22 (-0.30, -0.15) | -0.11 (-0.18, -0.03) | 0.30 (0.21, 0.40) | 0.16 (0.07, 0.26) |
| Animal naming | 7 625 | -0.52 (-0.58, -0.46) | -0.22 (-0.28, -0.16) | -0.21 (-0.29, -0.13) | -0.08 (-0.16, -0.01) | 0.25 (0.15, 0.34) | 0.10 (0.00, 0.19) |
| Letter cancellation speed | 7 412 | -0.24 (-0.30, -0.18) | -0.19 (-0.26, -0.13) | -0.13 (-0.20, -0.05) | -0.11 (-0.19, -0.03) | 0.09 (-0.01, 0.18) | 0.05 (-0.04, 0.15) |
| Delayed word recall | 7 623 | -0.53 (-0.59, -0.48) | -0.28 (-0.34, -0.22) | -0.26 (-0.34, -0.19) | -0.16 (-0.23, -0.08) | 0.27 (0.17, 0.36) | 0.14 (0.05, 0.24) |

Table S5. Associations between type of high school attended and health outcomes at age 46 (outcomes: average of three readings), comprehensive/other as reference group

| Outcomes at 46 years | N | Grammar; Z-score (95% CI) | | Private; Z-score (95% CI) | |
|---|---|---|---|---|---|
| | | Sex-adjusted models | Fully adjusted models | Sex-adjusted models | Fully adjusted models |
| Systolic blood pressure | 7 179 | 0.05 (-0.06, 0.17) | -0.01 (-0.13, 0.11) | 0.18 (0.10, 0.27) | 0.10 (0.01, 0.19) |
| Pulse | 7 177 | 0.17 (0.05, 0.29) | 0.09 (-0.04, 0.21) | 0.20 (0.11, 0.29) | 0.09 (-0.01, 0.19) |
| Grip strength | 7 146 | -0.02 (-0.10, 0.06) | -0.04 (-0.12, 0.04) | 0.02 (-0.04, 0.08) | -0.01 (-0.08, 0.05) |

Table S6. Associations between type of qualification/university attended and health outcomes at age 46 (outcomes: average of three readings), normal-status university as reference group

| Outcomes at 46 years | N | No qualification/GCSEs Z-score (95%CI) | | A-levels/diplomas Z-score (95%CI) | | Higher-status university Z-score (95%CI) | |
|---|---|---|---|---|---|---|---|
| | | Sex-adjusted models | Fully adjusted models | Sex-adjusted models | Fully adjusted models | Sex-adjusted models | Fully adjusted models |
| Systolic blood pressure | 6 828 | -0.16 (-0.22, -0.10) | -0.11 (-0.18, -0.05) | -0.13 (-0.21, -0.06) | -0.11 (-0.19, -0.03) | 0.04 (-0.06, 0.14) | 0.02 (-0.08, 0.12) |
| Pulse | 6 826 | -0.22 (-0.28, -0.16) | -0.15 (-0.22, -0.08) | -0.13 (-0.22, -0.05) | -0.10 (-0.19, -0.02) | 0.02 (-0.08, 0.12) | -0.01 (-0.11, 0.10) |
| Grip strength | 6 791 | 0.02 (-0.02, 0.06) | 0.06 (0.01, 0.11) | 0.02 (-0.03, 0.07) | 0.03 (-0.02, 0.09) | -0.02 (-0.08, 0.05) | -0.05 (-0.11, 0.02) |

Table S7. Associations between type of high school attended and health outcomes at age 46, models adjusted for sex and childhood cognitive factors (comprehensive and other as reference group)

| Outcomes at 46 years | N | Grammar Z-score (95% CI) | Private Z-score (95% CI) |
|---|---|---|---|
| **-Cardiometabolic** | | | |
| Body mass index | 7 068 | 0.05 (-0.07, 0.18) | 0.22 (0.12, 0.31) |
| Systolic blood pressure | 7 180 | 0.02 (-0.10, 0.13) | 0.15 (0.06, 0.23) |
| Pulse | 7 178 | 0.10 (-0.03, 0.22) | 0.13 (0.04, 0.22) |
| **-Physical function** | | | |
| Grip strength | 7 146 | -0.02 (-0.09, 0.06) | 0.01 (-0.05, 0.07) |
| Standing balance: eyes opened | 7 054 | -0.05 (-0.18, 0.07) | 0.02 (-0.07, 0.12) |
| Standing balance: eyes closed | 6 118 | 0.18 (0.05, 0.31) | 0.18 (0.09, 0.28) |
| **-Cognitive function** | | | |
| Immediate word recall | 8 052 | 0.07 (-0.04, 0.18) | 0.13 (0.05, 0.22) |
| Animal naming | 8 047 | 0.11 (0.00, 0.22) | 0.16 (0.07, 0.24) |
| Letter cancellation speed | 7 822 | 0.01 (-0.11, 0.13) | 0.15 (0.06, 0.24) |
| Delayed word recall | 8 046 | 0.11 (0.00, 0.22) | 0.12 (0.03, 0.20) |

Table S8. Associations between type of qualification/university attended and health outcomes at age 46, models adjusted for sex and childhood cognitive factors (normal status university as reference group)

| Outcomes at 46 years | N | No qualification/GCSEs Z-score (95%CI) | A-levels/diplomas Z-score (95%CI) | High-status university Z-score (95%CI) |
|---|---|---|---|---|
| **-Cardiometabolic** | | | | |
| Body mass index | 6 715 | -0.24 (-0.31, -0.17) | -0.22 (-0.31, -0.14) | 0.11 (0.01, 0.22) |
| Systolic blood pressure | 6 829 | -0.14 (-0.20, -0.07) | -0.12 (-0.20, -0.04) | 0.02 (-0.07, 0.12) |
| Pulse | 6 827 | -0.17 (-0.24, -0.11) | -0.11 (-0.19, -0.03) | 0.01 (-0.10, 0.11) |
| **-Physical function** | | | | |
| Grip strength | 6 791 | 0.05 (0.01, 0.09) | 0.03 (-0.02, 0.08) | -0.03 (-0.10, 0.03) |
| Standing balance: eyes opened | 6 698 | -0.10 (-0.17, -0.03) | -0.04 (-0.12, 0.04) | 0.01 (-0.10, 0.11) |
| Standing balance: eyes closed | 5 805 | -0.21 (-0.29, -0.14) | -0.18 (-0.27, -0.09) | 0.07 (-0.04, 0.17) |
| **-Cognitive function** | | | | |
| Immediate word recall | 7 630 | -0.26 (-0.32, -0.20) | -0.11 (-0.19, -0.04) | 0.18 (0.08, 0.27) |
| Animal naming | 7 625 | -0.25 (-0.31, -0.19) | -0.10 (-0.17, -0.02) | 0.12 (0.03, 0.21) |
| Letter cancellation speed | 7 412 | -0.20 (-0.27, -0.13) | -0.11 (-0.19, -0.03) | 0.07 (-0.03, 0.17) |
| Delayed word recall | 7 623 | -0.29 (-0.35, -0.23) | -0.16 (-0.23, -0.08) | 0.15 (0.06, 0.25) |

Table S9. Associations between type of high school attended and health outcomes at age 46, comprehensive and other as reference group.
(Participants with complete outcome data at age 46, N=5 726)

| Outcomes at 46 years | Grammar; Z-score (95% CI) | | Private; Z-score (95% CI) | |
|---|---|---|---|---|
| | Sex-adjusted models | Fully adjusted models | Sex-adjusted models | Fully adjusted models |
| Body mass index | 0.18 (0.06, 0.31) | 0.08 (-0.05, 0.20) | 0.28 (0.19, 0.37) | 0.14 (0.04, 0.23) |
| Systolic blood pressure | 0.05 (-0.08, 0.17) | -0.01 (-0.12, 0.13) | 0.17 (0.08, 0.26) | 0.10 (0.01, 0.20) |
| Pulse | 0.20 (0.07, 0.33) | 0.14 (0.01, 0.27) | 0.21 (0.11, 0.30) | 0.12 (-0.01, 0.22) |
| Grip strength | -0.00 (-0.08, 0.08) | -0.00 (-0.09, 0.08) | 0.00 (-0.06, 0.06) | -0.01 (-0.07, 0.06) |
| Standing balance: eyes opened* | - | - | - | - |
| Standing balance: eyes closed | 0.29 (0.15, 0.42) | 0.17 (0.03, 0.30) | 0.24 (0.15, 0.34) | 0.08 (-0.03, 0.18) |
| Immediate word recall | 0.38 (0.25, 0.51) | 0.03 (-0.10, 0.16) | 0.42 (0.32, 0.51) | 0.07 (-0.03, 0.17) |
| Animal naming | 0.38 (0.25, 0.51) | 0.04 (-0.09, 0.17) | 0.43 (0.33, 0.53) | 0.09 (-0.01, 0.19) |
| Letter cancellation speed | 0.11 (-0.02, 0.24) | -0.01 (-0.14, 0.12) | 0.23 (0.14, 0.33) | 0.10 (-0.00, 0.20) |
| Delayed word recall | 0.36 (0.23, 0.49) | 0.06 (-0.07, 0.18) | 0.37 (0.28, 0.46) | 0.07 (-0.03, 0.17) |

*models did not converge

Table S10. Associations between type of qualification/university attended and health outcomes at age 46, normal-status university as reference group.
(Participants with complete outcome data at age 46, N=5 431)

| Outcomes at 46 years | No qualification/GCSEs Z-score (95%CI) | | A-levels/diplomas Z-score (95%CI) | | Higher-status university Z-score (95%CI) | |
|---|---|---|---|---|---|---|
| | Sex-adjusted models | Fully adjusted models | Sex-adjusted models | Fully adjusted models | Sex-adjusted models | Fully adjusted models |
| Body mass index | -0.26 (-0.32, -0.19) | -0.19 (-0.26, -0.12) | -0.24 (-0.32, -0.16) | -0.20 (-0.29, -0.12) | 0.12 (0.02, 0.23) | 0.09 (-0.02, 0.19) |
| Systolic blood pressure | -0.16 (-0.22, -0.09) | -0.13 (-0.21, -0.06) | -0.14 (-0.22, -0.05) | -0.12 (-0.21, -0.04) | 0.02 (-0.09, 0.12) | 0.01 (-0.10, 0.11) |
| Pulse | -0.21 (-0.28, -0.14) | -0.16 (-0.23, -0.08) | -0.16 (-0.24, -0.07) | -0.13 (-0.21, -0.04) | 0.02 (-0.09, 0.12) | -0.01 (-0.12, 0.10) |
| Grip strength | 0.05 (0.01, 0.10) | 0.07 (0.03, 0.12) | 0.00 (-0.05, 0.06) | 0.01 (-0.04, 0.07) | -0.02 (-0.09, 0.05) | -0.04 (-0.10, 0.03) |
| Standing balance: eyes opened* | - | - | - | - | - | - |
| Standing balance: eyes closed | -0.27 (-0.34, -0.20) | -0.19 (-0.26, -0.11) | -0.21 (-0.30, -0.12) | -0.17 (-0.26, -0.08) | 0.07 (-0.04, 0.18) | 0.02 (-0.09, 0.13) |
| Immediate word recall | -0.49 (-0.56, -0.42) | -0.22 (-0.29, -0.15) | -0.21 (-0.30, -0.12) | -0.09 (-0.18, -0.01) | 0.31 (0.20, 0.41) | 0.18 (0.08, 0.28) |
| Animal naming | -0.48 (-0.55, -0.41) | -0.20 (-0.27, -0.13) | -0.19 (-0.28, -0.10) | -0.05 (-0.14, 0.03) | 0.24 (0.13, 0.35) | 0.11 (0.00, 0.21) |
| Letter cancellation speed | -0.24 (-0.31, -0.18) | -0.17 (-0.25, -0.10) | -0.14 (-0.23, -0.06) | -0.11 (-0.20, -0.02) | 0.10 (-0.01, 0.20) | 0.06 (-0.05, 0.16) |
| Delayed word recall | -0.49 (-0.55, -0.42) | -0.26 (-0.33, -0.18) | -0.26 (-0.34, -0.17) | -0.15 (-0.24, -0.07) | 0.25 (0.15, 0.35) | 0.15 (0.04, 0.25) |

*models did not converge

Figure S1. Flow chart on analytical sample selection

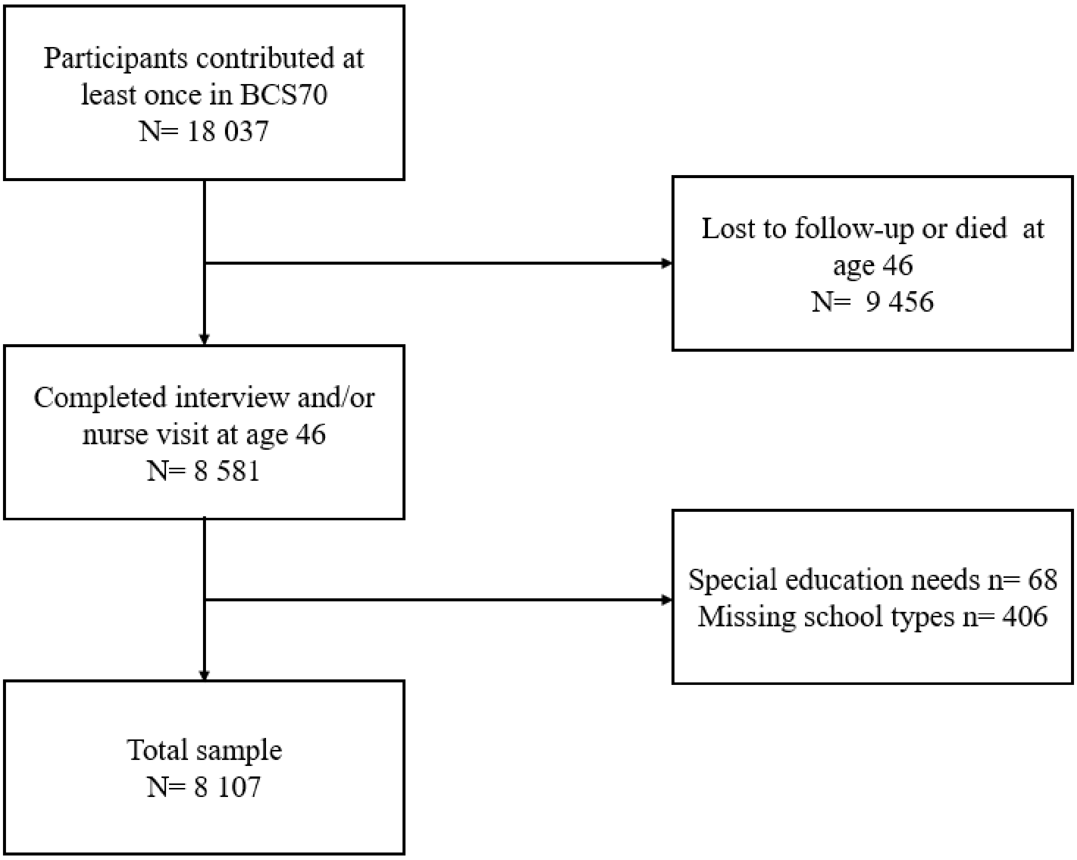

Figure S2. Cognitive score (percentile) at age 10 and rate of grammar/private school attendance

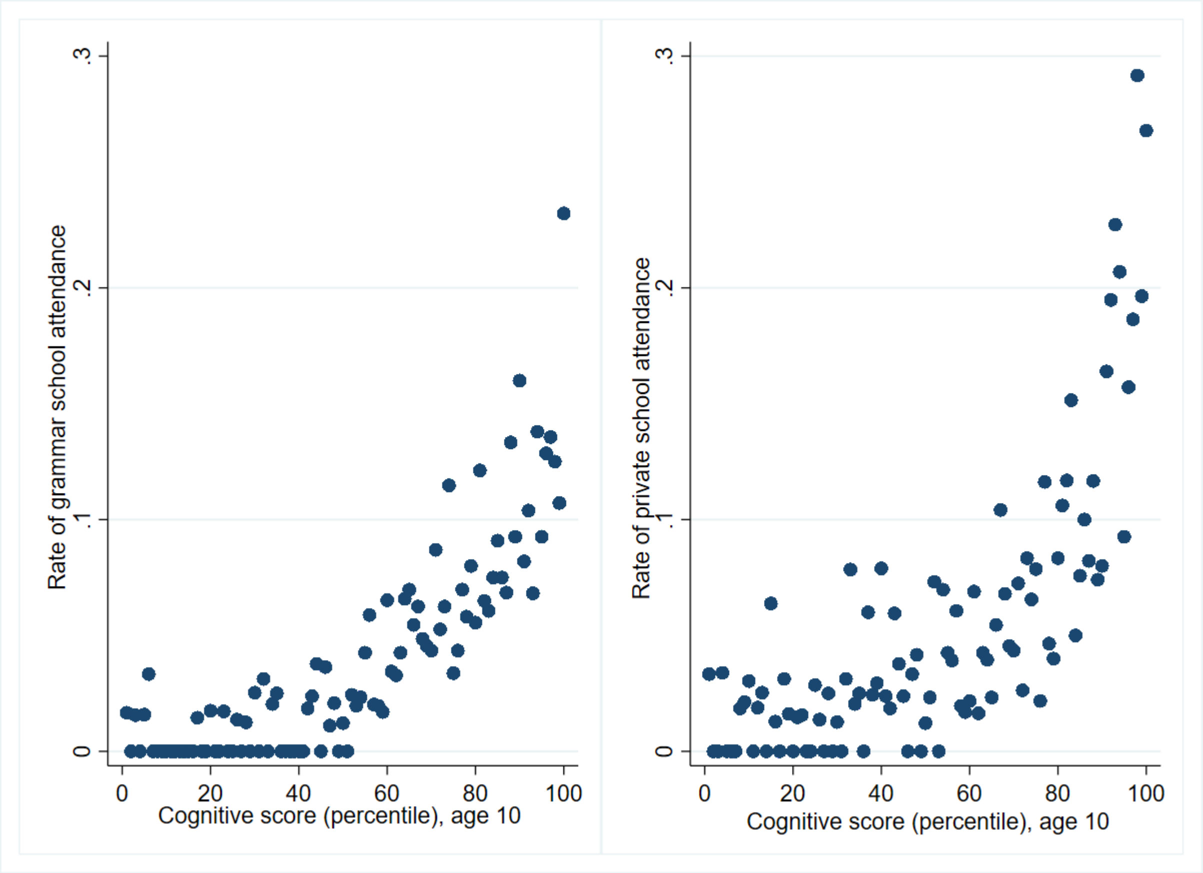
